# Determinants of anti-retroviral therapy adherence among adolescents living with HIV in the Kingdom of Eswatini

**DOI:** 10.1101/2023.09.25.23296111

**Authors:** Londiwe D. Hlophe, Constance S. Shumba, Diribsa T. Bedada, Peter S. Nyasulu

**Author notes:** Corresponding authors (LDH), (PSN).

## Abstract

**Background:** The success of antiretroviral therapy (ART) depends on a high level of adherence to the life-long therapy of 95% and above. However, in Eswatini, adolescents and young people aged 15 to 24 years, have poor ART adherence as evidenced by low viral load (VL) suppression rates (76% and 63% among female and males respectively) compared to the general population (96%). The wide age-band of 15 to 24 years in reporting viral suppression rates means that adolescent specific data is limited, and younger adolescents aged 10-14 years are excluded. This study explored the level of ART adherence and associated factors among adolescents living with HIV (ALHIV) aged 10 to 19 years on ART in Eswatini.

**Methods:** We performed a retrospective analysis of 911 medical records of ALHIV aged 10 to 19 years on ART for at least a month in Eswatini for the period 1^st^ January 2017 to 30^th^ September 2022. Bivariate logistic regression was fitted for each predictor variable. Missing values were imputed using multiple imputation by chained equation (MICE). Statistically significant (p-value ≤0.2) predictor variables were included in a multivariable logistic regression model. P-value ≤0.05 was used to declare statistical significance in the final regression model.

**Results:** ART adherence of 88.5% was recorded with higher adherence among males (88.9%) than females (87.8%). Hhohho region had highest adherence (90.7%) while Shiselweni region had lowest adherence (82%). Adherence was low among ALHIV with a non-suppressed baseline VL result (65.3%) and those assigned to multi-month ART model of care at ART initiation (66.7%). The Shiselweni region (OR 0.47; 95%CI 0.26-2.78), suppressed baseline VL result (OR 5.49; 95%CI 3.36-8,96) and assigned to the main-stream ART care model (OR 0.22; 95%CI 0.05-0.95) were statistically associated with ART adherence. In the multivariable regression analysis, only Shiselweni region and suppressed baseline VL result were statistically associated with ART adherence.

**Conclusion:** Eswatini ALHIV have a low ART adherence compared to general population. Shiselweni region is negatively associated with ART adherence among ALHIV while a suppressed baseline VL is positively associated with improved ART adherence. There is therefore a need to intensify interventions aimed at early ART initiation and intensive follow-up and support among ALHIV especially in the Shiselweni region.

## 1. Background

Globally, commendable strides have been made in attaining an optimum antiretroviral therapy (ART) adherence rate, and accordingly achieving viral load suppression (VLS) among people living with HIV (PLHIV). For instance, in the year 2021, 75% of PLHIV were on ART and 68% of these were virally suppressed [1,2]. In eastern and southern Africa which is home to 54% of all PLHIV globally, 79% of PLHIV were on ART, and of these, 78% were virally suppressed in 2021 [3,4].

ART adherence is defined as *“the extent to which a personal behaviour in taking HIV medication, attending scheduled clinic appointments, following a diet or certain behaviour corresponds to the treatment and care plan conjointly agreed between the person living with HIV and the health worker”* [5]. Optimum ART adherence, defined as 95% and above of correctly taking medication is a primary determinant of VLS [6]. Viral load suppression is viral load <1000 RNA copies per ml of blood and is critical in prevention of transmission of HIV to sero-negative partner, disease progression, opportunistic diseases, and AIDS-related deaths [5–7].

However, consistent with the evidence, globally, high ART uptake and VLS rates are only observed among the general population with adolescents reporting lower rates, yet adolescents account for 5% of all PLHIV [3,8]. The rates are even lower among ALHIV in sub-Sahara Africa (SSA) [1,3,9–11]. For instance, in 2021, VLS was only 30% among the 43% ALHIV on ART in SSA [12]. Furthermore, a systematic review including studies conducted between 2010 and 2022, reported an ART adherence rate of 65% and a VLS rate of 55% among ALHIV in SSA [13]. Similarly, studies conducted in South Africa, Kenya, Tanzania, and Uganda have reported even lower ART adherence rate ranging from 59% to 65% and VLS ranging from 22% to 73% [14–19]. Several factors associated with ART adherence have been identified among studies. These include age, sex, missing clinical appointment, presence of opportunistic diseases, duration on ART, ART regimen, family support, type of health facility, World Health Organization (WHO) clinical staging, VLS, and CD4-count [10,18,20–23].

The Kingdom of Eswatini formerly known as Swaziland has the highest Human Immuno-deficiency Virus (HIV) prevalence of 27% globally with an incidence of 1.4 among those 15 years and above [24]. Females are mostly affected in the country with an HIV prevalence of 32.5% compared to 20.4% among males [25]. Secondly, the HIV prevalence is disproportionally high at 7.3% among female adolescents aged 15 to 19 years compared to their male counterparts at 3.9% [25]. HIV prevention and management is critical among adolescents not only in Eswatini, but in the sub-Saharan Africa (SSA) region which is home to 90% of adolescents living with HIV (ALHIV) [3].

Eswatini is among the first countries globally to meet the 95-95-95 (95% people living with HIV knowing their HIV status, 95% of these on ART and 95% of those on ART viral load suppressed) UNAIDS global targets in the general population ahead of the year 2030 [26]. Viral load suppression (VLS) was reported to be 96% as of 2022 in the country, from 91% in 2017 [25,26]. This has been attributed to the uninterrupted access and availability of antiretroviral therapy (ART) among those testing positive to HIV and programmes aimed at supporting ART adherence [27].

However, these positive gains are only observed among the general population while ALHIV still present poor HIV treatment outcomes [28]. For instance, in 2017 Eswatini reported 89% of the people living with HIV (PLHIV) on ART with 91% of them virally suppressed. However, among ALHIV and young people, only 81.7% were on ART and 76.4% virally suppressed [25]. The discrepancies in viral load suppression rate among adolescents and general population, are an indication of poor HIV prevention and management, particularly poor ART adherence among ALHIV.

Unfortunately, there is limited HIV data on adolescents aged 10 to 19 years in Eswatini due to the wide description age-band of 15 to 24 years. Therefore, this study aimed to assess the level of ART adherence and associated factors among ALHIV aged 10 to 19 years on ART in Eswatini.

## 2. Methods

### 2.1 Study design and population

This was a retrospective study design which used data from the Eswatini Ministry of Health, Health Management Information System [HMIS]. Data extraction was done using a chart abstraction tool from the Client Management Information System (CMIS) for adolescents who accessed care and tested HIV positive between the periods of January 2017 and 30 September 2022. In 2017, Eswatini adopted and implemented the universal test and treat guidelines and the same year, the CMIS was rolled out in all the health facilities in the country [29]. The CMIS is a national electronic medical record system aimed at tracking and caring for patients visiting health facilities in the country. The system consolidates all the medical history of patients into one data set allowing for comprehensive health information of patients [29].

### 2.2 Inclusion and exclusion criteria

This study included ALHIV who were enrolled and initiated in ART at age 10–19 with either vertical or horizontal HIV infection from 1^st^ January 2017 to 30^th^ September 2022. Only ALHIV and had been on ART for at least a month were included in this study. ALHIV who transitioned from paediatric care were excluded from this study. Secondly, ALHIV whose records had missing details of latest or current viral load (VL) result in the CMIS database were excluded from the study. Current or latest VL result was defined as the last available VL result.

### 2.3 Ethical approval

Approval to carry out the study was sought from the Eswatini Health and Human Research Review Board of the Ministry of Health and a waiver of consent was obtained from the Board (Protocol Reference Number: EHHRRB103/2022). A waiver not to acquire participants’ informed consent was granted by the same board as this study involved secondary data analysis. Anonymity was maintained by de-identifying the data and assigning codes instead of names during the chart abstraction process.

### 2.4 Study setting

The data involved ALHIV enrolled in ART care from 1^st^ January 2017 to 30^th^ September 2022 in the Kingdom of Eswatini and registered in CMIS database. In 2015, adolescents accounted for 10% of all those in HIV treatment in the country [30]. Adolescents access care through a differential ART service delivery model which includes expert client offered by lay heath workers (LHWs) through Teen Clubs, multi-months (3 and 6 months) refills, as well as the mainstream model [31,32]. The expert client approach involves training LHWs on HIV treatment adherence, stigma and disclosure, and HIV linkage to care and communication and counselling skills [33,34]. The mainstream model also known as the standard of care model provides direct patient monitoring by a nurse or a doctor mostly for patients newly initiated to ART, with detectable VL, with ART adherence issues, or suspected treatment failure [35]. In 2021, there were 10,153 ALHIV in the country while there were 81 Teen Clubs with 3,742 adolescents aged 10 to 19 years as members. The country has 327 health facilities; one (1) national referral hospital, three (3) specialized hospitals, five (5) regional referral hospitals, five (5) health centres, six (6) public health units, 297 clinics of which 65 are specialized clinics, and nine (9) private hospitals [36]. These health facilities are distributed in the four regions (Hhohho, Manzini, Lubombo and Shiselweni) of the country with the national referral hospital in the Hhohho region.

### 2.5 Study sample

A total 3,420 ALHIV aged 10 to 19 were enrolled on ART care between 1^st^ January 2017 and 30th September 2022. Of these, 911 were included in the analysis as they met the inclusion criteria. Among the 911, some had missing data in one or more variables. We imputed the missing information using Multivariate Imputation by Chained Equations (MICE) for the variables we decided to include into the final multivariable regression model.

### 2.6 Measurements

All participants were followed for the period between January 2017 up until September 2022. Only ALHIV enrolled on ART between 1^st^ January 2017 and September 2022 were included in this study. Our primary outcome was ART adherence defined as 95% of correctly taking ART which was estimated using the viral load suppression rate where viral load suppression was defined as viral load <1000 RNA copies per ml of blood, a standard VLS measurement used in Eswatini [37].

Predictor variables included: demographic and social factors (sex, age, region), HIV factors (age at diagnosis, baseline VL result, latest viral load result, ART start date, time on ART, model of ART care at initiation, latest or current model of ART care, regimen, CD4 count result, and WHO clinical staging), and facility factors (facility ownership). Baseline VL result was defined as the first VL result after a positive HIV diagnosis. Facility ownership included government, non-government, private and mission.

### 2.7 Statistical analysis

The dataset in Excel was exported to Stata for the data analysis, and the analysis was done using the Stata statistical software, release 16, College Station, TX, Stata Corp LLC.

Data analyses were conducted at descriptive, bivariate, and multivariable levels. Continuous variables (age, years at HIV diagnosis and years on ART) were presented as means with standard deviations and categorical variables were presented as frequencies and percentages. Differences in characteristics between ALHIV who were adherent versus those who were non-adherent were assessed using the chi-square test. Associations between ART adherent and independent variables were examined using the Pearson’ chi-square test.

Variables that were significantly associated with ART adherence at bivariate analysis at significance level of ≤ 0.2 were selected and included in the multivariable regression model. Age and sex were included despite their lack statistical significance. Due to high missing data in some of the variables, a Multiple Imputation by Chained Equation (MICE) was conducted to impute the missing values.

Backward elimination was used in building the final model whereby variables which did not contribute at p-value <0.05 were eliminated and a new model fitted. All variables that were significant at p<0.05 were kept in the final multivariable logistic regression model.

## 3. Results

### 3.1 Socio-demographic characteristics of participants

A total of 3420 medical records were reviewed of which only 911 were included in the analysis. Six-hundred-and-six (66.6%) adolescents were in the age group 15 to 19 years and 155 (60%) were females. The mean age of the ALHIV and on ART was 16.3 (SD 3.3). Majority (355; 39%) of the adolescents were from the Hhohho region while only 133 (15%) were from the Shiselweni region (Table 1).

**Table 1:**
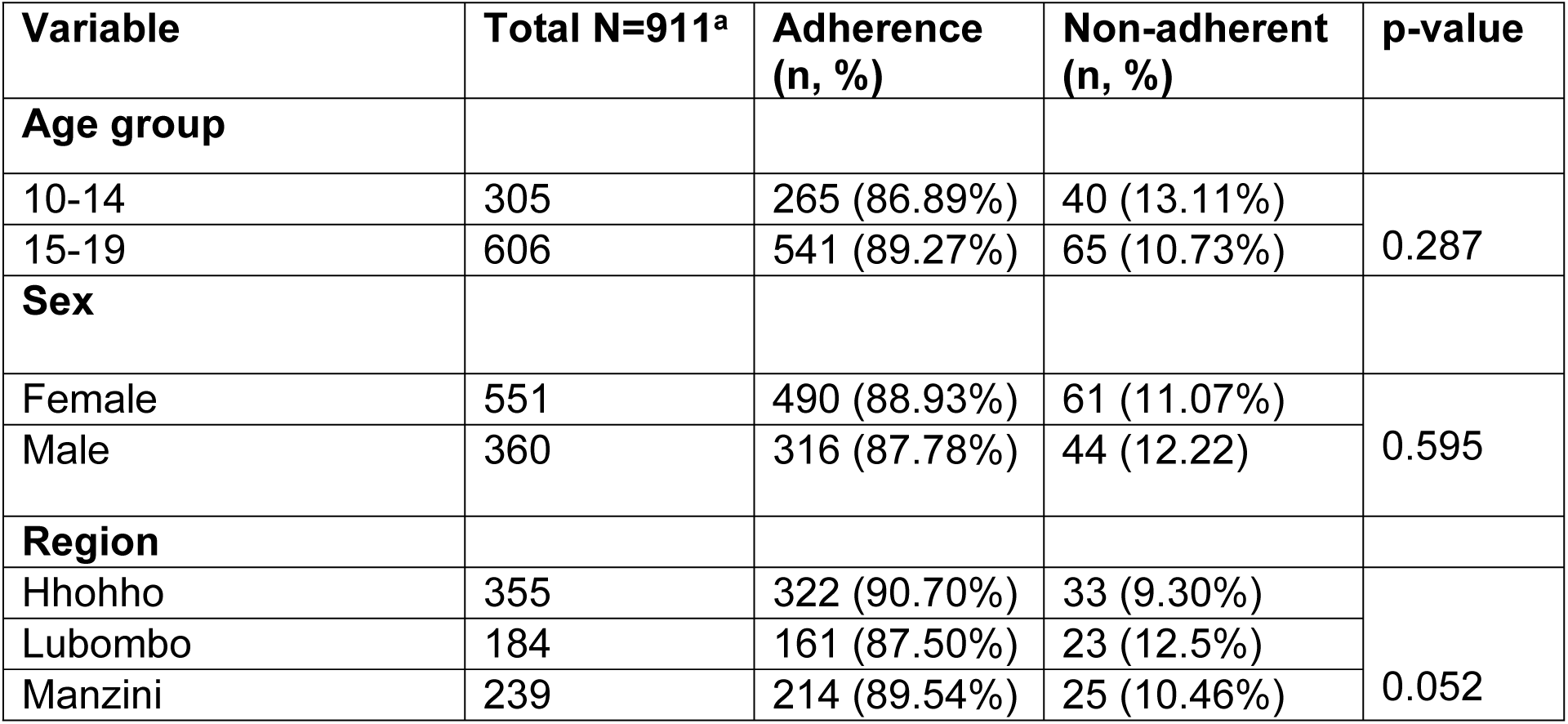
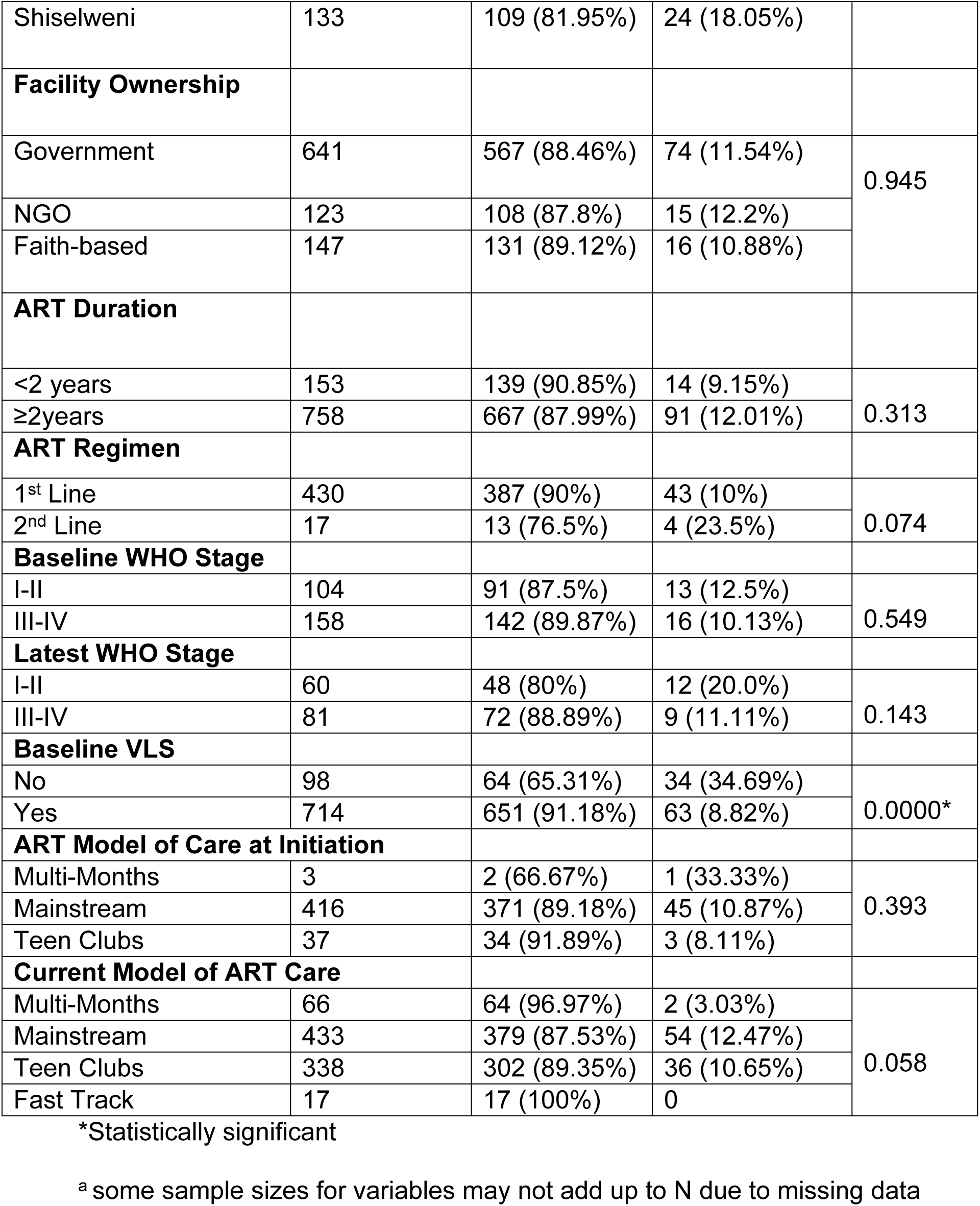
Characteristics of study participants stratified by ART adherence.

### 3.2 Clinical and Treatment-Related Characteristics

The average age at diagnosis of adolescents was 14 years (SD 4.2). Most adolescents (355; 70%) were enrolled under government owned health facilities while only 123 (13.5%) were enrolled in both private and non-government health facilities. Government health facilities include both military and non-military health facilities. Among adolescents with a recorded CD4 count results, 158 (60%) were on either World Health Organization (WHO) clinical stage three or stage 4 as a baseline CD4 count result. As of 2022 (latest CD4 result), only 81 out of 141 adolescents with CD4 count result were on either of these stages. Seven-hundred-and-fifty-eight (83%) of the adolescents had been on ART for 2 years and above. At baseline, 714 (88%) adolescents were virally load suppressed (Table 1). Overall, ART adherence was 88.5% among adolescents. The age group 15 to 19 years had higher adherence (89.3%) compared to 10-14 years (86.9%). Hhohho region recorded 90.7% ART adherence followed by Manzini (89.5%), Lubombo (87.50%), while Shiselweni had the lowest adherence rate of 81.95% (Table 1).

### 3.3 Factors Associated with ART Adherence among ALHIV

In the bivariate logistic regression analysis, Shiselweni region, viral load suppressed at initiation and mainstream at current ART care model were statistically significant predictors and were fitted in the multivariable logistic regression model (Table 2). In the multivariable regression model (Table 3), only Shiselweni and baseline or initial viral load suppressed were predictors of ART adherence.

**Table 2:** Factors associated with ART adherence among ALHIV.

| Variable | OR | p-value | 95% CI |
| --- | --- | --- | --- |
| **Age** |  |  |  |
| 10-14 | Ref |  |  |
| 15-19 | 1.26 | 0.287 | 0.83-1.91 |
| **Sex** |  |  |  |
| Female | Ref |  |  |
| Male | 0.89 | 0.595 | 0.59-1.35 |
| **Region** |  |  |  |
| Hhohho | Ref |  |  |
| Lubombo | 0.72 | 0.249 | 0.41-1.26 |
| Manzini | 0.88 | 0.639 | 0.51-1.52 |
| Shiselweni | 0.47 | 0.008* | 0.26-2.78 |
| **Facility Ownership** |  |  |  |
| Government | Ref |  |  |
| Non-government and Private | 0.92 | 0.789 | 0.51-1.67 |
| Faith-based | 1.07 | 0.820 | 0.60-1.89 |
| **ART Duration** |  |  |  |
| <2 years | Ref |  |  |
| ≥2 years | 0.74 | 0.315 | 0.41-1.33 |
| **Regimen** |  |  |  |
| 1<sup>st</sup> Line | Ref |  |  |
| 2<sup>nd</sup> Line | 0.36 | 0.086 | 0.11-1.16 |
| **Baseline VLS** |  |  |  |
| No | Ref |  |  |
| Yes | 5.49 | 0.000* | 3.36-8.96 |
| **Baseline WHO stage** |  |  |  |
| I-II | Ref |  |  |
| III-IV | 1.27 | 0.550 | 0.58-2.78 |
| **Latest WHO stage** |  |  |  |
| I-II | Ref |  |  |
| III-IV | 2 | 0.148 | 0.78-5.11 |
| **ART Model of Care at Initiation** |  |  |  |
| Multi-Month | Ref |  |  |
| Mainstream | 4.12 | 0.251 | 0.37-46.37 |
| Teen Club | 5.67 | 0.204 | 0.39-82.24 |
| **Current ART Model of Care** |  |  |  |
| Multi-Month | Ref |  |  |
| Mainstream | 0.22 | 0.038* | 0.05-0.92 |
| Teen Club | 0.26 | 0.070 | 0.06-1.12 |

**Table 3:** Multivariate Logistic Regression showing factors associated with ART adherence among ALHIV.

| Variable | OR | p-value | 95% CI |
| --- | --- | --- | --- |
| **Age group** |  |  |  |
| 10-14 | Ref |  |  |
| 15-19 | 1.49 | 0.231 | 0.81-2.54 |
| **Sex** |  |  |  |
| Female | Ref |  |  |
| Male | 1.35 | 0.632 | 0.72-1.98 |
| **Region** |  |  |  |
| Hhohho | Ref |  |  |
| Lubombo | 0.86 | 0.412 | 0.52-1.73 |
| Manzini | 0.90 | 0.512 | 0.63-1.79 |
| Shiselweni | 0.54 | 0.029* | 0.17-0.96 |
| **Baseline VLS** |  |  |  |
| No | Ref |  |  |
| Yes | 6.22 | 0.001* | 4.13-10.56 |
| **Current ART Model of Care** |  |  |  |
| Multi-Month | Ref |  |  |
| Mainstream | 1.23 | 0.661 | 0.67-2.23 |
| Teen Clubs | 0.94 | 0.623 | 0.58-2.14 |
| Constant | 8.22 | 0.031 | 2.65-24.812 |

\*Statistically significant

The odds of adolescents from Shiselweni region were negatively associated (OR=0.54, 95%CI: 0.17-0.96) with ART adherence than adolescents from the other region of the country. The odds of adolescents with a suppressed baseline viral load were (OR=6.22, 95% CI: 4.13-10.56) almost six-fold likely to adhere to ART compared to those who were non-virally load suppressed (Table 3).

## 4. Discussion

The study revealed the determinants of ART adherence among ALHIV in the Kingdom of Eswatini. In this study, the mean age was 16.3 (SD 3.3) years with more females (551, 60%) compared to males (360, 40%). On average, adolescents were initiated on ART at the age of 14 years (SD 4.2). The study discovered that overall, ART adherence among ALHIV was sub-optimal at 88.5% which is lower than the 95% ART adherence rate required for suppressed viral load. When compared to other studies, ART adherence among Swati adolescents was higher than in studies conducted among ALHIV in South Africa (65%), Kenya (76%) and Zimbabwe (28%) while comparable with studies conducted in Nigeria (85%), Uganda (80%), Cameroon (83%) [10,16,22,38–40]. These discrepancies among the countries could be due to the differences in outcome measurements, settings, sample sizes, and study designs as explained in a study conducted in Ethiopia among children on ART [21].

Nevertheless, an ART adherence rate below 95% is associated with unsuppressed viral load which is further associated with opportunistic infections, drug resistance and subsequently death [7]. For instance, in two studies conducted in South Africa, lower ART adherence rate was statistically associated with increased opportunistic infections and a two-fold (OR 1.98) likelihood of a detectable viral load [41,42]. Another study conducted among ALHIV in South Africa reported that poor ART adherence was also associated with virologic failure [43]. A study conducted in Liberia also revealed that poor ART adherence was associated with loss to follow up and experiences of drug side effects [44].

Secondly, the study findings indicate that Shiselweni region is a predictor of poor ART adherence compared to the other regions. This finding is not comparable to the national findings which indicate that generally the Shiselweni region has the highest HIV prevalence (26.5%) compared to the other regions but not statistically associated with either viral load suppression or ART adherence [27]. The Swaziland HIV Incidence measurement survey (SHIMS II) also reported that the different HIV prevalence rates in the four regions of the country had no significant difference [25]. However, findings of this study have revealed that there is an association between ART adherence and region among ALHIV. This could be because previous studies were conducted among the general population or adult population while this study was conducted among adolescent age group. Studies conducted in the Shiselweni region revealed that the universal test and treat (UTT) HIV program has been associated with low retention to care partly due to status non-acceptance by those tested positive and other factors such as transport costs and distance to health facilities [45–47]. The Shiselweni region is ranked the poorest region in the country with high food and water insecurities [46,48]. According to the Eswatini operational plan of 2020, the population of the Shiselweni region is hard-to-reach with limited access to health services and HIV prevention, control, and management services such as HIV testing and VL testing since it is dynamic and very evanescent because of the recent influx and growth of textile factories, and it harbours the main truck route and border gate [48]. The region is therefore a hot spot for at-risk population [48]. The delayed initiation of ART due to low testing in the Shiselweni region has been associated with poor ART outcomes such as low viral load suppression among those who are eventually tested in the region [46,47].

According to studies conducted in Uganda and South Africa, retention to care was a positive predictor of ART adherence while a study conducted in South Africa also revealed that distance to health facilitator was statistically associated with ART adherence [18,49]. Studies aimed at improving the financial capacity of ALHIV have reported a positive association with retention to care and ART adherence [50–54]. This has been facilitated by financial incentives aimed at assisting ALHIV with money for honouring their hospital appointments, curbing food insecurities among others. For instance, a study conducted in Uganda which was aimed at evaluating the efficacy of economic incentive in improving ART outcomes such as viral load suppression and adherence among ALHIV, presented a ten-fold increase among those receiving the economic incentive compared to standard of care [52]. However, status acceptability has been reported to be associated with retention to care and ART adherence. In studies conducted in Zambia, Tanzania and South Africa, early status disclosure among adolescents was associated with early acceptability of status and thus treatment adherence [55–57]. Additionally, a study conducted by Cluver *et al.* (2015) revealed that adolescents who had their status disclosed early were three times likely to be retained in care and thus improved ART adherence [55]. However, other studies have reported that adolescents whose status were revealed early were most likely to miss medication doses due to treatment fatigue [49,58]. These contradicting findings are justified by Ankrah *et al.* (2016) because of lack of support system among ALHIV especially older adolescents as care-giver responsibility reduces and adolescents are at a self-discovery stage [59]. Nevertheless, early treatment to initiation and adherence to the treatment are critical in improving ART outcomes. WHO guidelines have established a positive association between early treatment initiation and improved ART outcomes such as viral load suppression which are associated with optimum ART adherence among those on ART [6]. Early initiation to ART is facilitated by the availability of health services promoting HIV testing for early diagnosis and subsequent treatment through the UTT approach [6,18].

Lastly, the study revealed that baseline viral load suppression is a positive predictor for ART adherence as revealed in other studies. Numerous studies have reported the association between VLS and ART adherence. High viral load suppression is positively associated with optimum ART adherence while non-viral load suppression is a predictor for poor ART adherence [60–67]. For instance, in a study conducted in Kenya, adolescents and children with a suppressed viral load were mostly likely to adhere to treatment compared to those with a high viral load at ART initiation [68]. Furthermore, the UTT has been based on scientifically proven evidence that early initiation to ART is effective in ensuring positive ART outcome such as reduced AIDS related morbidity and mortality [69–73]. The positive ART outcomes are enabled by the low viral load which associated with ART adherence.

The strength of our study is that the results demonstrate the current ART adherence level and key associated factors (demographic and social factors, HIV factors, and facility factors) among ALHIV aged 10 to 19 years, a population group normally reported with young adults and paediatrics. Secondly, since the study used the national dataset, the study results can extrapolate the current ART adherence situation in Eswatini specifically among ALHIV. Additionally, since secondary data was used, recall and reporting bias were eliminated. Also, the data collection process was informed by professionalism and expertise, and not associated with research thus was not subject to interview bias. Lastly, being the only study conducted among adolescents within this age group, our study can contribute to informing policies, plans, and service delivery strategies for preventing and controlling poor ART adherence among ALHIV in Eswatini.

Nonetheless, some limitations should be considered when interpreting the findings of this study. Firstly, this was a retrospective study that used secondary data for analysis that was not intentionally collected for this study, and therefore limited the study in evaluating other predictors of ART adherence which were not covered in the records. Secondly, being a secondary dataset made the data prone to missing variables due to incomplete documentation. However, we are confident that the available data provide true estimation as it was captured by qualified health professionals.

## 5. Conclusions

In conclusion, our results demonstrate that ART adherence is sub-optimum among ALHIV in Eswatini despite the positive progress in attaining the 95-95-95 UNAIDS 2030 target by the general population. It is critical to better understand why adolescents have a sub-optimum adherence rate and device strategies aimed at improving ART adherence among ALHIV in Eswatini. These results highlight an urgent need to innovate strategies aimed at improving early ART initiation and intensive follow-up and support among ALHIV especially in the Shiselweni region.

## Data Availability

All relevant data are within the manuscript.

## 6. List of abbreviations

AIDS: Acquired Immune Deficiency Syndrome
OR: Odds ratio
ART: Anti-Retroviral Therapy
HIV: Human Immunodeficiency Virus
VL: Viral Load
VLS: Viral Load suppression
ALHIV: Adolescents Living with HIV
PLHIV: People Living with HIV
WHO: World Health Organization
CMIS: Client Management Information System
SSA: sub-Sahara Africa
LHW: Lay Health Worker
UTT: Universal Test and Treat

## 7. Declaration

### Ethics approval and consent to participate

Ethical approval for conducting the study was sought from the Eswatini Health and Human Research Review Board of the Ministry of Health and a waiver of consent was obtained from the Board since the study used secondary data (Protocol Reference Number: EHHRRB103/2022).

### Consent of publication

The Eswatini Health and Human Research Review Board of the Ministry of Health granted the consent for publication of the findings of the study.

### Availability of data and materials

Data underlying the findings of the study are included in the paper.

### Authors’ contribution

Conceptualization: Londiwe D. Hlophe, Peter S. Nyasulu, Constance S. Shumba

Dara curation: Londiwe D. Hlophe, Diribsa T. Bedada

Formal analysis: Londiwe D. Hlophe, Diribsa T. Bedada

Investigation: Londiwe D. Hlophe

Methodology: Londiwe D. Hlophe, Diribsa T. Bedada, Constance S. Shumba, Peter S. Nyasulu

Writing-original draft preparation: Londiwe D. Hlophe

Writing-review and editing: Londiwe D. Hlophe, Diribsa T. Bedada, Constance S. Shumba, Peter S. Nyasulu

## Acknowledgement

The authors would like to thank the management and staff from the Ministry of Health; Health Management Information System for the support provided during the duration of the study. Special gratitude is also forwarded to Dr. Garikai Chemhaka for his valuable support throughout the study.

